# Neoadjuvant Bevacizumab in Newly Diagnosed, Surgically Resectable Glioblastoma: A Systematic Review and Meta-Analysis of Survival and Functional Outcomes

**DOI:** 10.1101/2025.10.03.25337250

**Authors:** Farzan Fahim, Farbod Tabasi Kakhki, Fatemeh Sadat Hosseini Khajouei, Aysan Valinejad Qanati, Maryam Babazadeh, Roozbeh Tavanaei, Melika Hajimohammadebrahim-Ketabforoush, Sayeh Oveisi, Saeed Oraee-Yazdani, Alireza Zali

## Abstract

**Background:** Glioblastoma (GBM) remains one of the most aggressive primary brain tumors, with limited survival despite maximal safe resection and chemoradiotherapy. Neoadjuvant bevacizumab (BEV) has been proposed to reduce peritumoral edema, improve functional status, and potentially enhance progression-free survival (PFS). However, its survival benefit in newly diagnosed, surgically resectable GBM remains unclear. Therefore, this systematic review and meta-analysis aimed to synthesize the available evidence regarding the survival and functional outcomes associated with neoadjuvant BEV in this population.

**Methods:** Following PROSPERO registration (CRD420251078761), we searched PubMed, Embase, Scopus, Web of Science, and Cochrane Library up to July 20, 2025, without language restrictions. Eligible studies evaluated bevacizumab administered before surgery either as monotherapy or in combination with other standard treatments such as temozolomide or radiotherapy. Combination regimens were allowed because several clinical studies have investigated bevacizumab as part of multimodal perioperative treatment strategies rather than as an isolated intervention.Primary outcomes were overall survival (OS) and PFS; secondary outcomes included Karnofsky Performance Status (KPS), steroid use, radiological response, and biomarkers. Data were pooled using a random-effects model.

**Results:** Ten studies (2 randomized controlled trials, 4 non-randomized clinical trials, and 4 cohort studies) met the inclusion criteria; four studies (n=751 participants) provided sufficient data for quantitative meta-analysis. Pooled HR for OS was 0.72 (95% CI: 0.42–1.25, p=0.246) and for PFS was 0.72 (95% CI: 0.42–1.22, p=0.220), both with low heterogeneity (I^2^=0%). Functional outcomes were assessed qualitatively because quantitative pooling was not feasible due to heterogeneity Available evidence suggested trends toward improved Karnofsky Performance Status (KPS) and reduced steroid dependence; however, the certainty of evidence was low.

**Conclusions:** Neoadjuvant BEV in resectable GBM does not significantly improve OS or PFS but may offer symptomatic and functional benefits. Current evidence is limited by small sample sizes, heterogeneous protocols, and low methodological quality. Well-designed multicenter RCTs are warranted.

## Introduction

Glioblastoma (GBM) is the most common and aggressive primary malignant brain tumor in adults, accounting for approximately 50–60% of all malignant gliomas, with an annual incidence of 3–4 cases per 100,000 population and a median overall survival (OS) of only 14–16 months despite standard treatment (1-3).Standard-of-care therapy for newly diagnosed, surgically resectable GBM includes maximal safe resection followed by concurrent radiotherapy with temozolomide (TMZ) and adjuvant TMZ (the Stupp protocol)(4). However, even with optimal resection, postoperative neurological deterioration, persistent peritumoral edema, and seizure or steroid dependence remain common, adversely affecting both quality of life and functional independence (5, 6).

Bevacizumab (BEV), a humanized monoclonal antibody targeting vascular endothelial growth factor (VEGF), has demonstrated potent anti-angiogenic effects, reduction of peritumoral vasogenic edema, and rapid radiologic responses in GBM (7-9). While its FDA approval is restricted to recurrent GBM based on radiographic response rates, multiple investigations have explored BEV in the adjuvant and recurrent settings, showing consistent improvements in progression-free survival (PFS) but no definitive OS benefit(10-12). In the neoadjuvant context (administration before surgical resection), BEV offers several theoretical advantages: improved tumor oxygenation, decreased vascular permeability, reduced intracranial pressure, and potentially safer, more extensive resections due to diminished edema (13-15). However, concerns have also been raised regarding the perioperative safety of bevacizumab. Because VEGF inhibition may impair angiogenesis and tissue repair, preoperative administration of BEV has been associated with potential risks such as delayed wound healing, increased bleeding tendency, and postoperative complications. These concerns have led to recommendations for an appropriate interval between the last bevacizumab dose and surgical resection in order to minimize perioperative risks.

Despite these potential benefits, evidence for neoadjuvant BEV in newly diagnosed, resectable GBM remains sparse and methodologically heterogeneous. Existing studies vary in dosing schedule (single versus multiple preoperative doses), concurrent therapies (BEV with or without TMZ and/or preoperative radiotherapy), and the interval between BEV administration and surgery (16, 17).Furthermore, although early reports suggest improvements in neurological and functional measures (particularly Karnofsky Performance Status (KPS) preservation, cognitive function, and steroid discontinuation),these outcomes have rarely been synthesized systematically or meta-analyzed(13, 18, 19). Given that maintaining KPS ≥70 is one of the strongest independent prognostic factors for survival in GBM(20), the functional impact of BEV may be as clinically relevant as its survival effects.

There is currently no comprehensive systematic review and meta-analysis exclusively evaluating neoadjuvant BEV in newly diagnosed, surgically resectable GBM, integrating both survival endpoints (OS, PFS) and functional/neurological outcomes. In addition, no prior synthesis has combined clinical results with imaging, histopathological, and biomarker findings to provide a multi-dimensional picture of BEV’s perioperative effects. Addressing this knowledge gap is timely, given the emerging interest in perioperative systemic therapies in neuro-oncology and ongoing debates regarding cost-effectiveness and patient selection based on predictive biomarkers (21-23).

Therefore, the present study systematically reviews and quantitatively synthesizes the available evidence from clinical trials and cohort studies on neoadjuvant BEV in newly diagnosed GBM patients eligible for surgical resection, with a dual focus on (1) survival outcomes (OS, PFS) and (2) functional and radiological endpoints, while also summarizing biological correlates where available. This approach aims to clarify the potential role of neoadjuvant BEV in contemporary GBM management and to identify gaps requiring future targeted research.

### Methodology

This systematic review and meta-analysis was registered in the International Prospective Register of Systematic Reviews (PROSPERO) to ensure methodological transparency and to avoid duplication. The protocol is publicly available in PROSPERO 2025 (CRD420251078761). The most recent version (Version 3.0, published 23 August 2025) includes amendments to the types of studies considered eligible for inclusion and introduces additional restrictions to the eligibility criteria. The literature search was completed on July 20, 2025. The PROSPERO record was subsequently updated (Version 3.0, August 23, 2025) to clarify eligible study designs and refine the predefined eligibility criteria. These amendments did not modify the research question, target population, intervention, or outcomes of interest. Instead, they aimed to improve methodological clarity and ensure that only studies evaluating preoperative bevacizumab in newly diagnosed, surgically resectable glioblastoma were included. All eligibility criteria were finalized before data extraction and were applied independently by reviewers during full-text screening using the Rayyan platform.

### Search Strategy

A comprehensive set of keywords was used to search for and collect relevant studies in PubMed, Scopus, Web of Science, Embase, and the Cochrane Library. No limitation of date and language was applied. The keywords included: “bevacizumab”, “chemotherapy, adjuvant”, “radiotherapy, adjuvant”, “adjuvants, pharmaceutic”, “neoadjuvant therapy”, “radiation”, “temozolomide”, “glioblastoma”, “glioblastoma multiforme”, “GBM”, “grade IV glioma”, “high-grade glioma”, and “Avastin”. Only articles published (including online-first publications) inception to 20 July 2025 were considered. The complete search strategies for all databases are provided in Supplementary File 2.

### Inclusion and Exclusion Criteria

Studies were eligible for inclusion if they involved adult patients (≥18 years) with histologically confirmed glioblastoma, including both newly diagnosed and resectable cases. The intervention of interest was bevacizumab (BEV), administered either as monotherapy, as neoadjuvant therapy, or in combination with other modalities such as temozolomide (TMZ) and radiotherapy (RT). For the purpose of this review, neoadjuvant bevacizumab (BEV) was defined as bevacizumab administered before surgical resection in patients with newly diagnosed, surgically resectable glioblastoma. Eligible studies were required to include at least one preoperative BEV administration prior to tumor resection. However, the reviewed literature demonstrated variability in how neoadjuvant therapy was operationalized, including differences in the number of preoperative cycles, concomitant therapies, and whether bevacizumab was continued after surgery as maintenance or administered again at recurrence. Consequently, while all included studies incorporated a preoperative BEV component, the broader treatment strategies sometimes extended beyond a strictly neoadjuvant design. Control groups differed across the included studies and generally reflected the standard-of-care treatment used in each trial. In randomized studies, control arms typically consisted of standard chemoradiotherapy with radiotherapy plus temozolomide (RT+TMZ) or placebo-controlled regimens without bevacizumab. In several cohort or non-randomized studies, the comparator group received conventional treatment strategies such as temozolomide-based therapy or surgery followed by standard chemoradiotherapy without bevacizumab. Consequently, the pooled hazard ratios should be interpreted as comparisons between treatment strategies that incorporated a preoperative bevacizumab component versus similar standard-of-care approaches without bevacizumab.

Eligible studies were required to report at least one relevant outcome. Primary efficacy outcomes included overall survival (OS), progression-free survival (PFS), and hazard ratios (HR) for survival with 95% confidence intervals (CI). Secondary and exploratory outcomes included median OS and PFS, overall response rates, and biomarker expression changes such as microvessel density (MVD), MIB-1 index, and Nestin-positive cells. Eligible study designs comprised randomized controlled trials (RCTs), non-randomized clinical trials, and cohort studies (prospective or retrospective) published in peer-reviewed journals.

Studies were excluded if they involved pediatric patients (<18 years old) or patients with brain tumor types other than histologically confirmed glioblastoma, such as astrocytoma, oligodendroglioma, or brain stem glioma, or if the diagnosis of glioblastoma was not confirmed. Also, studies including patients with unresectable or recurrent high-grade glioma was excluded. Studies were also excluded if the intervention did not involve bevacizumab (BEV) in any form (monotherapy, neoadjuvant, or combination therapy). For the specific analysis of neoadjuvant efficacy, studies in which bevacizumab was not administered in a preoperative setting were excluded. In terms of outcomes, studies that did not report any of the predefined primary or secondary outcomes (overall survival, progression-free survival, hazard ratios, median values, overall response rates, biomarker data, or adverse events) or those with non-extractable or unusable data were excluded. With respect to study design, we excluded non-primary research publications (such as reviews, editorials, letters, and conference abstracts without sufficient data), preclinical studies including animal models or in-vitro experiments, duplicate reports of the same study population, and case series.

### Screening and Study Selection

A comprehensive search of the aforementioned databases yielded a total of 9358 records. After removing duplicate entries, as well as reviews, books, speeches, conference abstracts, editorials, commentaries, and expert opinions, 3,988 studies remained for title and abstract screening. The title and abstract screening phase was independently performed by two reviewers (FT, MB) using the Rayyan platform to facilitate blinded and efficient screening, based on predefined PICOS-based inclusion and exclusion criteria. Any disagreements were resolved through discussion, and unresolved conflicts were adjudicated by a third reviewer (FF). Full texts of potentially eligible studies were then retrieved for detailed assessment. The full-text screening was independently conducted by two other reviewers (AV, FS), with any conflicts again resolved by consensus and, when needed, by arbitration from the third reviewer (FF).

The study selection process is illustrated in Figure 1 using the PRISMA 2020 flow diagram. This diagram summarizes the identification, screening, eligibility assessment, and final inclusion of studies in the systematic review.

**Figure 1.**
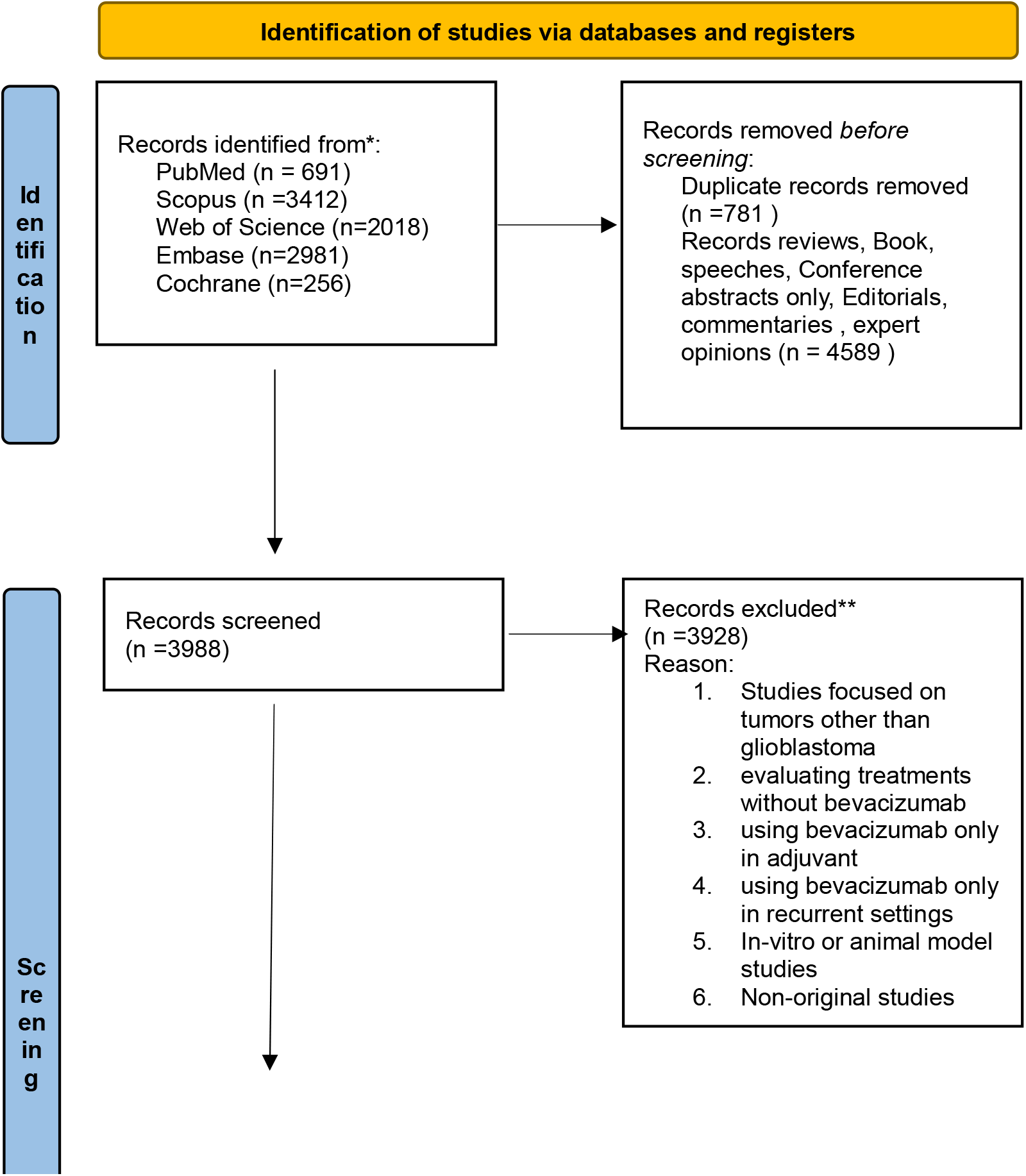
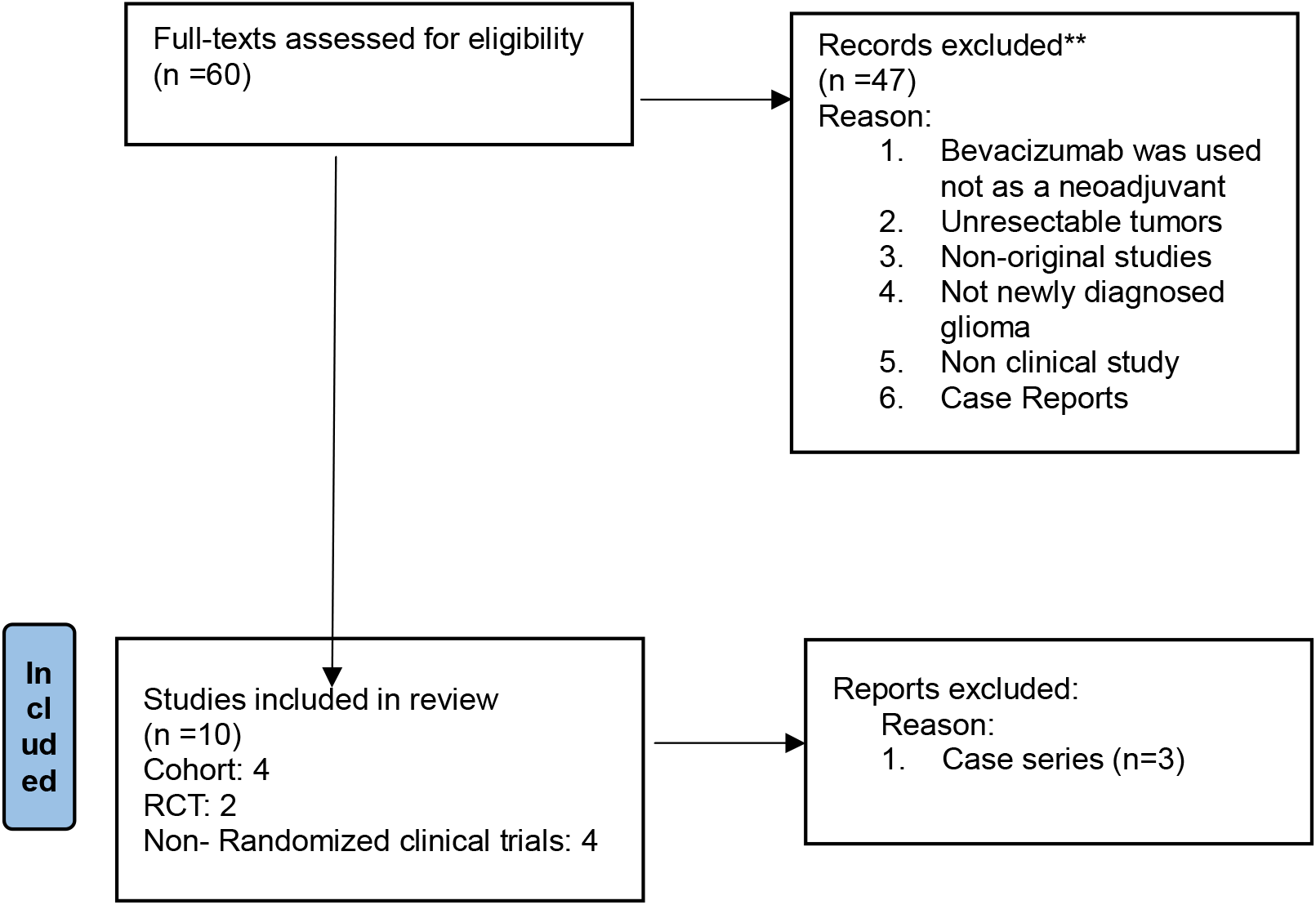
PRISMA 2020 flow diagram

### Data Extraction

Data extraction was performed independently by two reviewers (FS and AV) using a predefined standardized form. Extracted variables included study characteristics, patient demographics, intervention details, comparator treatments, survival outcomes (OS and PFS), hazard ratios, adverse events, and biomarker data where available. Discrepancies were resolved through discussion with a third reviewer (FF).

### Quality Assessment

Methodological quality was assessed independently by two reviewers (FT and MB) using the Joanna Briggs Institute (JBI) Critical Appraisal Checklists appropriate for each study design. Disagreements were resolved through discussion, and final judgments were based on consensus across predefined checklist domains.

### Data Synthesis and Statistical Analysis

For quantitative synthesis, effect sizes for time-to-event outcomes (OS and PFS) were expressed as hazard ratios (HRs) with 95% confidence intervals (CIs). For each eligible study, the natural logarithm of the HR (logHR) and its standard error (SE) were calculated from reported HRs and 95% CIs, or reconstructed from Kaplan–Meier curves when necessary (25). The meta-analysis was conducted using the meta package in R (R Foundation for Statistical Computing, Vienna, Austria). Pooled estimates were obtained with a random-effects model to account for between-study variability, with the restricted maximum-likelihood (REML) method used to estimate τ^2^ (between-study variance). Between-study heterogeneity was assessed using Cochran’s Q test (p<0.10 indicating significance) and quantified by the I^2^ statistic. Pooled logHRs were exponentiated back to HRs for ease of interpretation. Publication bias was evaluated using funnel plots. All statistical tests were two-sided, with p < 0.05 considered statistically.

### Certainty of evidence (GRADE approach)

The certainty of evidence for the primary and secondary outcomes was assessed using the GRADE framework. Two reviewers independently evaluated the evidence across five domains: risk of bias, inconsistency, indirectness, imprecision, and publication bias. Overall, the certainty of evidence was rated as low for overall survival (OS), progression-free survival (PFS), and functional outcomes, and very low for radiological and biomarker outcomes due to methodological limitations, small sample sizes, and heterogeneity across studies. A detailed Summary of Findings table and full GRADE assessments are provided in Supplementary File 3.

The characteristics and outcomes of studies evaluating neoadjuvant bevacizumab in glioblastoma are summarized in Table 1. Although ten studies met the eligibility criteria for the qualitative synthesis, not all studies reported data for every outcome domain. Consequently, the number of studies contributing to each outcome analysis varied depending on the availability of relevant data (e.g., functional outcomes, radiological findings, molecular analyses, or safety outcomes).

**Table 1.**
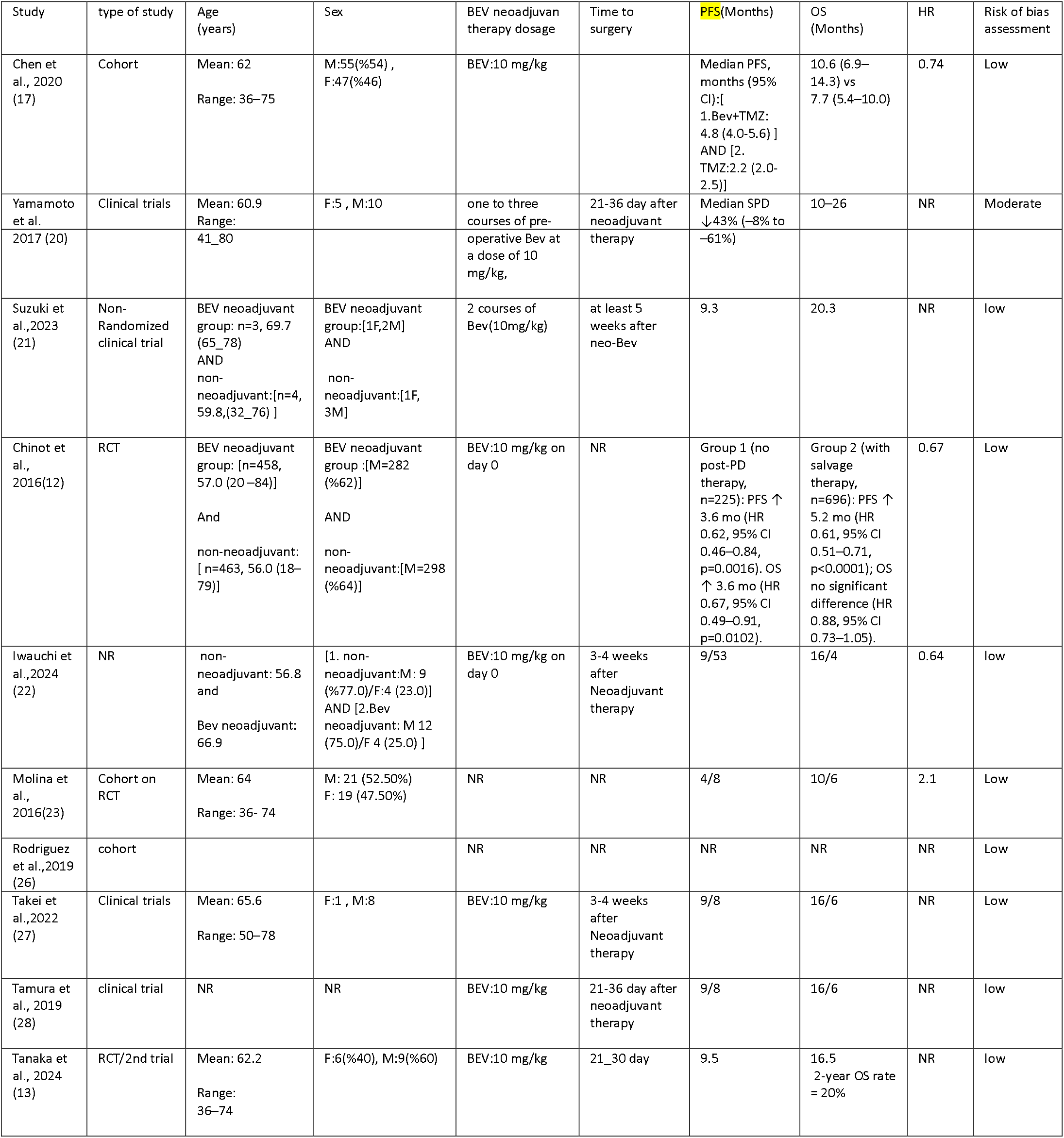
Characteristics and outcomes of studies evaluating neoadjuvant bevacizumab in patients with newly diagnosed, surgically resectable glioblastoma.

Age and sex data are reported as presented in the original studies. When information was not available in the source articles, it is indicated as NR (not reported)

## Result

### Neoadjuvant bevacizumab treatment protocol

As summarized in Table 2, six studies provided detailed descriptions of the neoadjuvant bevacizumab treatment protocols, including the number of preoperative cycles, concomitant therapies, and the interval between the final BEV infusion and surgery. Chen et⍰al., ⍰2020⍰ (17): BEV (days⍰1 & 15) combined with oral temozolomide (TMZ, 85⍰mg/m^2^/day, days1⍰–21) for two 28-day cycles before surgery. Postoperatively, BEV was permitted for up to six maintenance cycles. Yamamoto et⍰al., ⍰2017⍰ (20): One to three BEV doses preoperatively; surgery performed 21–36⍰days after the final infusion. Suzuki et⍰al., ⍰2023⍰ (21): Two BEV doses followed by RT (40–60⍰Gy) and continuous TMZ (75⍰mg/m^2^, 42⍰days), with surgery ≥⍰5⍰weeks later; maintenance BEV+TMZ thereafter. Takei et⍰al., ⍰2022⍰ (27): Single-dose BEV; surgery in 3– 4⍰weeks; BEV+TMZ resumed at recurrence and continued (mean⍰= ⍰16.1 cycles). Tanaka et⍰al., ⍰2024⍰ (13): Single BEV+TMZ course; surgery in 3–4⍰weeks; no postoperative BEV maintenance. Chinot et⍰al., ⍰2016⍰ (12): BEV started preoperatively and continued as long-term maintenance.

**Table 2.** Neoadjuvant bevacizumab protocols.

| Study (Year) | BEV Dose | Cycles | Concomitant Therapy | Surgery Interval from bevacizumab | Maintenance BEV | References |
| --- | --- | --- | --- | --- | --- | --- |
| Chen, 2020 | 10 mg/kg * | 2 | TMZ | Post-cycle | ≤ 6 cycles | (17) |
| Yamamoto, 2017 | * | 1-3 | none | 21–36 days | None | (20) |
| Suzuki, 2023 | * | 2 | RT + TMZ | ≥ 5 weeks | BEV+TMZ | (21) |
| Takei, 2022 | * | 1 | None; TMZ+BEV at recurrence | 3–4 weeks | Yes in recurrence | (27) |
| Tanaka, 2024 | * | 1 | TMZ | 3–4 weeks | No | (13) |
| Chinot, 2016 | * | NR | None | NR | Yes | (12) |

\*: same dose / NR: not reported / BEV: bevacizumab / TMZ: temozolomide/ RT: radiotherapy/

These differences, particularly in timing to surgery and whether BEV continued postoperatively or at recurrence, created significant *clinical* heterogeneity, despite similar dosing.

Although all included studies administered bevacizumab prior to surgical resection, the definition of “neoadjuvant” varied across trials. In some studies, BEV was used strictly as a short preoperative intervention before surgery, whereas in others it formed part of a broader perioperative treatment strategy that included continuation of BEV after surgery or at tumor recurrence. For example, Chen et⍰al. allowed postoperative maintenance BEV for up to six cycles, Suzuki et⍰al. continued BEV in combination with temozolomide after surgery, and Chinot et⍰al. incorporated long-term BEV administration as part of the treatment course. In contrast, Tanaka et⍰al. administered a single preoperative BEV course without postoperative maintenance. These differences indicate that the interventions ranged from purely neoadjuvant regimens to perioperative or extended treatment In addition to differences in the number of neoadjuvant cycles, important variation existed in how bevacizumab was combined with other therapeutic modalities. Some studies administered BEV together with temozolomide prior to surgery (e.g., Chen 2020; Tanaka 2024), whereas others used BEV without concomitant systemic therapy in the preoperative period (e.g., Yamamoto 2017). In Suzuki 2023, strategies. BEV was incorporated into a multimodal strategy that included radiotherapy and continuous temozolomide prior to surgery. Furthermore, postoperative treatment strategies differed substantially. Chen 2020 allowed maintenance BEV for up to six cycles after surgery, Suzuki 2023 continued BEV combined with temozolomide, and Chinot 2016 implemented long-term BEV administration as part of the treatment course. In contrast, Tanaka 2024 did not include postoperative BEV maintenance. These differences indicate that neoadjuvant bevacizumab was frequently embedded within broader perioperative treatment strategies rather than functioning as an isolated intervention.

### Primary Outcomes: Overall Survival (OS) and Progression-Free Survival (PFS)

As presented in Table 3, meta-analysis of four eligible studies (n = 751) demonstrated no statistically significant pooled effect of neoadjuvant bevacizumab (BEV) on either PFS or OS: PFS: HR = 0.7175 (95% CI: 0.4224–1.2188), p = 0.2195, I^2^ = 0% OS: HR = 0.7249 (95% CI: 0.4211–1.2481), p = 0.2459, I^2^ = 0%

**Table 3.** Pooled survival outcomes.

| Outcome | No. studies (n patients) | Pooled HR (95% CI) | p-value | I² (%) | Certainty (GRADE) | References |
| --- | --- | --- | --- | --- | --- | --- |
| PFS | 4 (751) | 0.7175 (0.4224–1.2188) | 0.2195 | 0 | Low | (17, 23) |
| OS | 4 (751) | 0.7249 (0.4211–1.2481) | 0.2459 | 0 | Low | * |

\*: same reference / PFS: Progression-Free Survival / OS: Overall Survival

**Table 4.**
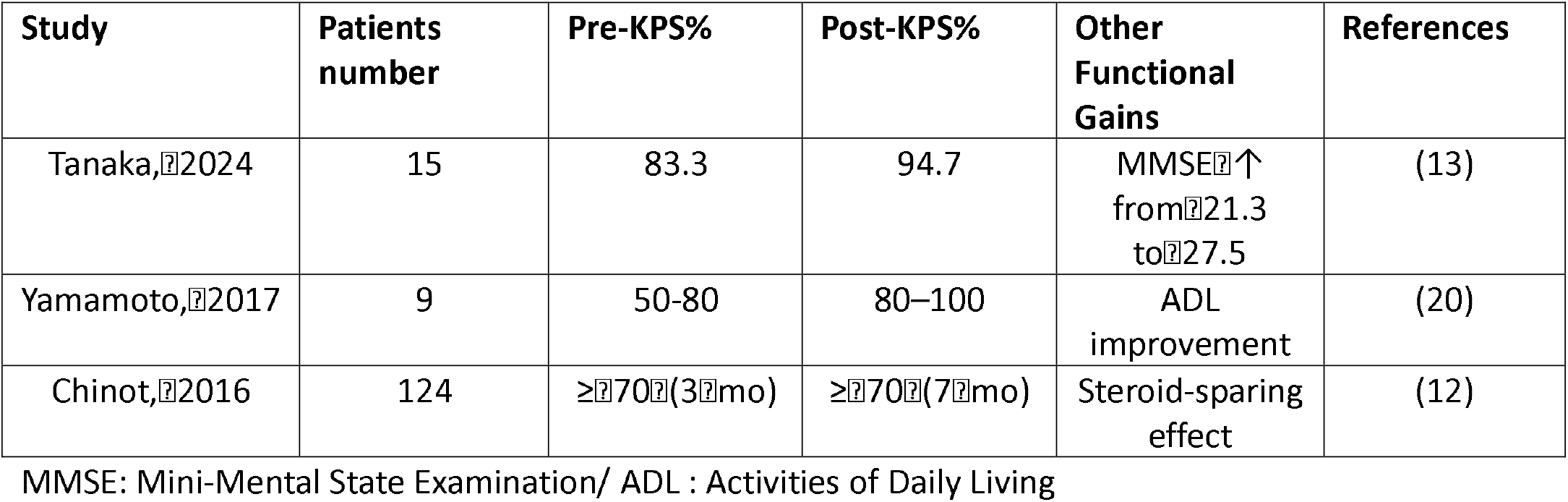
Functional outcome measures.

**Table 5.** Radiological changes with neoadjuvant BEV.

| Study | Imaging Modality | Main Findings | References |
| --- | --- | --- | --- |
| Tanaka, 2024 | MRI T1CE/FLAIR | Tumor vol ↓36.2% / edema ↓54.0% | (13) |
| Takei, 2022 | MRI T1Gd/FLAIR | Tumor vol ↓38% / edema ↓54% | (27) |
| Suzuki, 2023 | FMISO-PET | HV ↓ ($p=0.03$) – transient effect | (21) |
| Rodriguez, 2019 | MRI multimodal | ↑ detection speed of progression | (26) |
| Molina, 2016 | MRI volumetry | Low necrosis → ↑ OS benefit | (23) |

MRI :Magnetic Resonance Imaging /T1CE :T1-weighted Contrast-Enhanced sequence /FLAIR :Fluid-Attenuated Inversion Recovery /T1Gd :T1-weighted Gadolinium-enhanced sequence/ FMISO-PET :<sup>18</sup>F-fluoromisonidazole Positron Emission Tomography/ HV :Hypoxic Volume/ OS :Overall Survival

**Table 6.** Histopathological/molecular effect.

| Study | Main Findings | References |
| --- | --- | --- |
| Suzuki, 2023 | ↓ CA9, PD-L1, CD34; temp. hypoxia/stemness modulation | (21) |
| Takei, 2022 | Pseudo-papillary structures post-BEV; CA9/HIF-1α spatial variation | (27) |
| Yamamoto, 2017 | ↓ MVD, ↓ stemness; hypoxia rebound at recurrence | (20) |
| Tamura, 2019 | ↓ PD-L1/Tregs; ↑ cytotoxic T-cells | (28) |
| Chen, 2020 | ↓ vascular density/ERG; vessel remodeling | (17) |

CA9 :Carbonic Anhydrase IX (hypoxia-associated enzyme) /PD-L1 :Programmed Death-Ligand<sup>1</sup> (immune checkpoint ligand) /CD34 :Cluster of Differentiation<sup>34</sup> (endothelial cell marker)/HV :Hypoxic Volume /HIF-1α : Hypoxia-Inducible Factor<sup>1</sup>-alpha (transcription factor induced by low oxygen) /MVD :Microvessel Density /FOXM1 : Forkhead Box<sup>1</sup>M1 (transcription factor linked to cell proliferation and stemness)/Tregs :Regulatory<sup>1</sup>T cells (immunosuppressive lymphocyte subset)/PD-1:Programmed Cell Death Protein<sup>1</sup> (immune checkpoint receptor) / CD3:Cluster of Differentiation<sup>3</sup> (pan-T-cell marker) /CD8 :Cluster of Differentiation<sup>8</sup> (cytotoxic<sup>1</sup>T-cell marker) / CD163 :Cluster of Differentiation<sup>163</sup> (marker of M2-type macrophages) /ERG :ETS-Related Gene (endothelial transcription factor) /Nestin :Intermediate filament protein expressed in neural stem/progenitor cells /MIB-1/Ki-67 : Proliferation index antigen /VEGF :Vascular Endothelial Growth Factor /VEGFR1 :Vascular Endothelial Growth Factor Receptor<sup>1</sup>

**Table 7.**
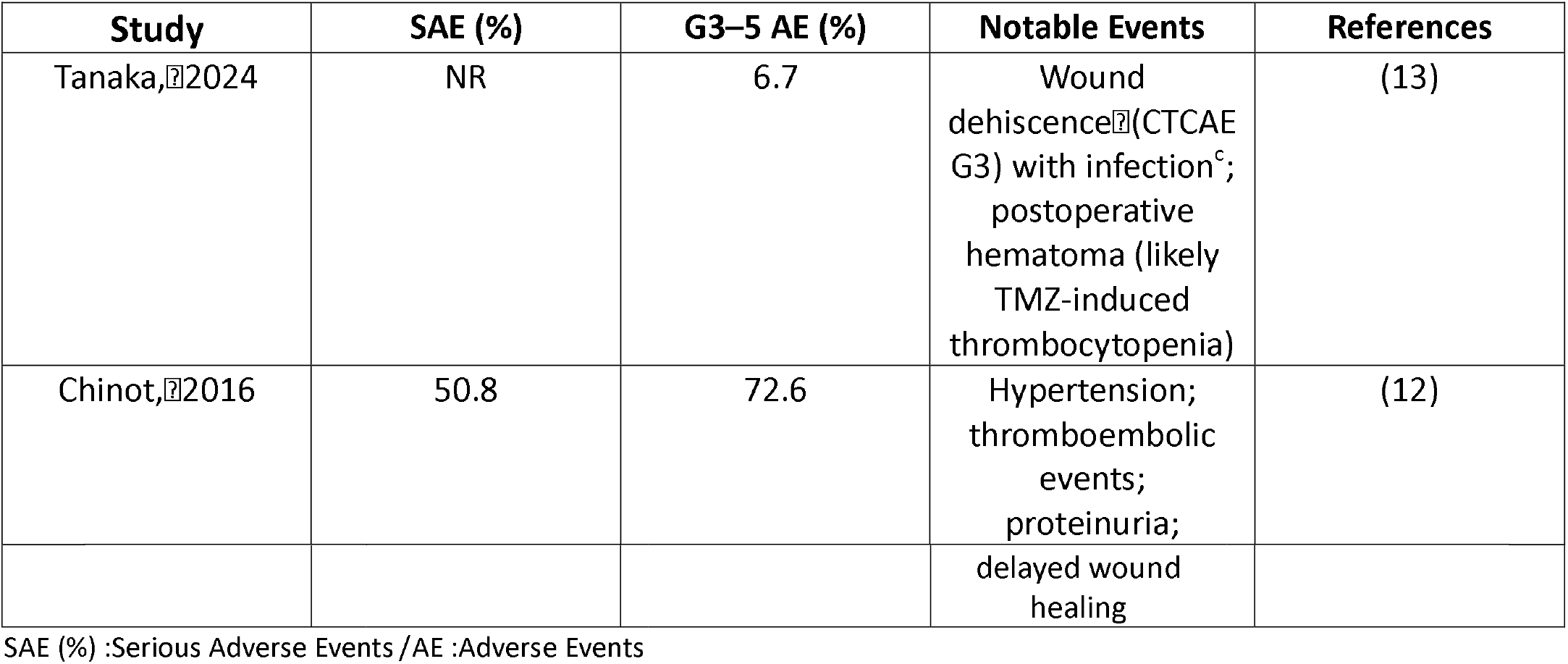
Safety profile.

The very low I^2^ indicates minimal statistical heterogeneity, whereas clinical heterogeneity was substantial due to differences in dosing schedules, combination with temozolomide ± preoperative radiotherapy, and timing of surgery.

In RCTs (Molina 2016 (23), Chen 2020 (17), Chinot 2016 (12)), numerical improvements in PFS occurred but did not reach significance in pooled analysis. In smaller single-arm trials (Tanaka 2024 (13), Takei 2022 (27)), median OS was ≈ 16 months and median PFS ≈ 9 months, but absence of control arms limits conclusions.

Sensitivity and publication bias: Funnel plots suggested no strong asymmetry. Egger’s test was not applied (< 10 studies/outcome). Leave-one-out analyses showed no excessive influence by any single study; pooled HRs changed by < 5% in all iterations.

### Neurological and Functional Outcomes

Functional outcome data were reported in three studies (Tanaka 2024, Yamamoto 2017, and Chinot 2016), primarily evaluating changes in Karnofsky Performance Status (KPS), cognitive function, or steroid use. Tanaka et al., 2024 (13): KPS rose from 83.3 ± 7.8 to 94.7 ± 4.5 (p < 0.0024); Mini-Mental State Examination (MMSE) from 21.3 to 27.5 (p < 0.0026). Yamamoto et al., 2017 (22): KPS improved from 50–80 to 80–100 in all patients (n=9). Chinot et al., 2016 (12): Median KPS ≥ 70 was 7 months with BEV vs 3 months placebo; corticosteroid discontinuation 70.6% vs 47.5%.

Functional outcomes were reported in three studies (Tanaka 2024, Yamamoto 2017, and Chinot 2016) and are summarized in Table 5. In Tanaka 2024, mean Karnofsky Performance Status (KPS) improved from 83.3 before treatment to 94.7 following neoadjuvant bevacizumab. Cognitive performance measured by the Mini-Mental State Examination (MMSE) increased from 21.3 to 27.5 after therapy. Yamamoto 2017 reported improvement in KPS categories from 50–80 before treatment to 80–100 following one to three preoperative BEV administrations, accompanied by improvements in activities of daily living. In the randomized study by Chinot 2016, the proportion of patients maintaining KPS ≥ 70 was sustained for a longer period in the BEV group (7 months) compared with the control group (3 months), and higher rates of steroid discontinuation were observed.

### Radiological Outcomes

Radiological outcomes were reported in five studies (Tanaka 2024, Takei 2022, Suzuki 2023, Rodriguez 2019, and Molina 2016), mainly assessing changes in tumor volume, peritumoral edema, or imaging biomarkers. Tanaka, 2024 (13) : Mean reduction: −36.2% (T1CE), −54.0% (FLAIR); larger T1CE reductions trended toward better OS. Takei, 2022 (27): Median reduction: −38% (T1Gd) and −54% (FLAIR). Suzuki, 2023 (21): FMISO-PET: HV ↓ significantly (p = 0.03), but changes transient. Rodriguez, 2019 (26): Multimodal MRI advanced detection of progression (∼6% earlier). Molina, 2016 (23): Lower pre-treatment necrotic volumes predicted OS benefit in BEV arm.

Radiological evaluations consistently demonstrated reductions in tumor burden and peritumoral edema following neoadjuvant BEV administration. In Tanaka 2024, mean tumor volume decreased by 36.2% on contrast-enhanced T1-weighted MRI (T1CE), while FLAIR-defined edema volume decreased by 54.0%. Similarly, Takei 2022 reported median reductions of 38% in gadolinium-enhanced tumor volume and 54% in FLAIR edema volume. Functional imaging data from Suzuki 2023 using FMISO-PET demonstrated a significant reduction in hypoxic volume (p = 0.03), although the effect appeared transient. Rodriguez 2019 reported that multimodal MRI enabled earlier detection of tumor progression, while Molina 2016 identified lower baseline necrotic volume as a potential predictor of overall survival benefit in BEV-treated patients

### Histopathological and Molecular Findings

Histopathological and molecular analyses were reported in five studies (Suzuki 2023, Takei 2022, Yamamoto 2017, Tamura 2019, and Chen 2020), evaluating vascular density, hypoxia-related markers, immune checkpoint molecules, and tumor microenvironment changes following neoadjuvant bevacizumab.Suzuki, 2023 (21): decrease C A9, PD-L1, CD163; decrease CD34; modulated FOXM1. Takei, 2022 (27): Pseudo-papillary structures at recurrence; differential CA9/HIF-1α expression. Yamamoto, 2017 (20): decrease MVD, decrease nestin+ stem-like cells; hypoxia rebounded at recurrence; decrease VEGF/VEGFR1. Tamura, 2019 (28): decrease PD-L1, Foxp3+, PD-1+; increase CD8+/CD3+ T-cell infiltration. Chen, 2020 (17): decrease CD34, ERG; vascular remodeling.

Five studies reported histopathological or molecular changes associated with neoadjuvant BEV exposure. Several studies demonstrated reductions in markers of angiogenesis and hypoxia. Suzuki 2023 observed decreased expression of CA9, PD-L1, and CD34, along with modulation of FOXM1 signaling. Takei 2022 reported pseudo-papillary structures in recurrent tumors with spatial variation in CA9 and HIF-1α expression. Yamamoto 2017 described reductions in microvessel density and nestin-positive stem-like cells, although hypoxia markers increased again at recurrence alongside VEGF and VEGFR1 upregulation. Tamura 2019 reported immunological changes including decreased PD-L1 expression and regulatory T cells with increased CD8^+^ and CD3^+^ T-cell infiltration. Chen 2020 documented decreased vascular density and endothelial remodeling marked by reduced CD34 and ERG expression.

### Safety and Adverse Events

Quantitative safety data were available from only two studies (Tanaka 2024 and Chinot 2016). Other included studies either reported adverse events qualitatively or did not provide sufficient numerical safety data for structured comparison. Tanaka, 2024 (13): No intraoperative hemorrhage; 1 wound dehiscence G3 (car-mustine-related). Chinot, 2016 (12): SAE: 50.8% vs 36.0% (control); G3–5 AE: 72.6% vs 63.0%.

Common: hypertension, thromboembolism, delayed wound healing.

Quantitative safety data were reported in two studies. In Tanaka 2024, no intraoperative hemorrhage was observed, and one patient developed grade 3 wound dehiscence with infection. In Chinot 2016, serious adverse events occurred in 50.8% of patients receiving BEV compared with 36.0% in the control group, and grade 3–5 adverse events were reported in 72.6% versus 63.0%, respectively. The most frequently reported complications included hypertension, thromboembolic events, proteinuria, and delayed wound healing. Safety reporting was inconsistent across other included studies, limiting the ability to perform pooled safety analysis.

### Cost-Effectiveness

Chen, 2020 (21): BEV+TMZ exceeded China’s willingness-to-pay threshold over 10 years; cost driven by drug price; biomarker-guided use recommended for economic viability

## 7. Analysis of Progression-Free Survival (PFS) and Overall Survival (OS)

A total of four studies comprising 751 participants were included in the meta-analysis. Individual study HRs for progression-free survival (PFS) ranged from 0.62 to 1.9. For overall survival (OS), individual study HRs ranged from 0.64 to 2.1. The pooled analysis using a random-effects model showed an HR of 0.7175 (95% CI, 0.4224–1.2188, p = 0.2195) for PFS and an HR of 0.7249 (95% CI, 0.4211–1.2481, p = 0.2459) for OS. There was low heterogeneity among studies for both OS and PFS (I^2^ = 0.0%). A forest plot illustrating individual study estimates and the overall pooled hazard ratio is presented in Figures 2 and 3. Despite substantial clinical heterogeneity in protocols, statistical heterogeneity was minimal (I^2^=0%). This may reflect the small number of studies and wide confidence intervals, and should not be interpreted as clinical homogeneity.

**Figure 2.**
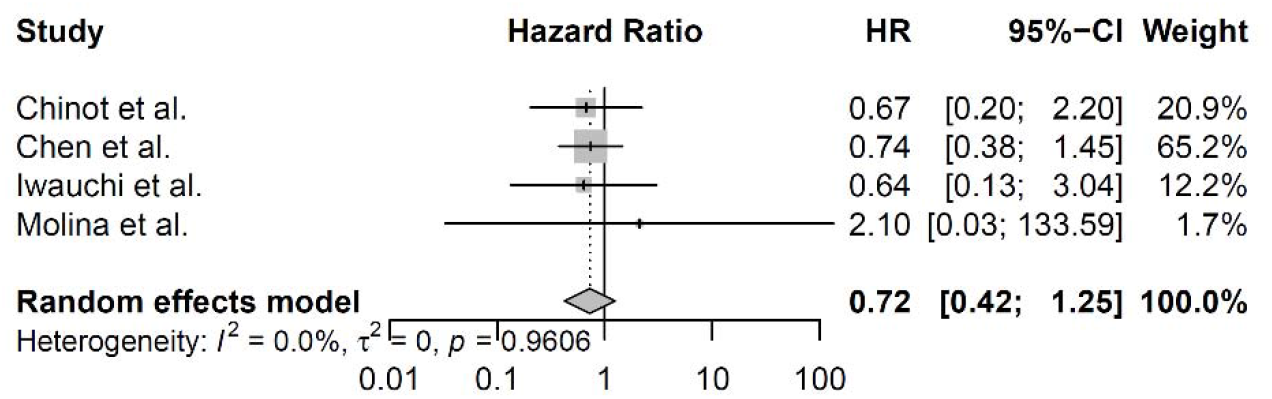
Forest plot of Overall Survival (OS).

**Figure 3.**
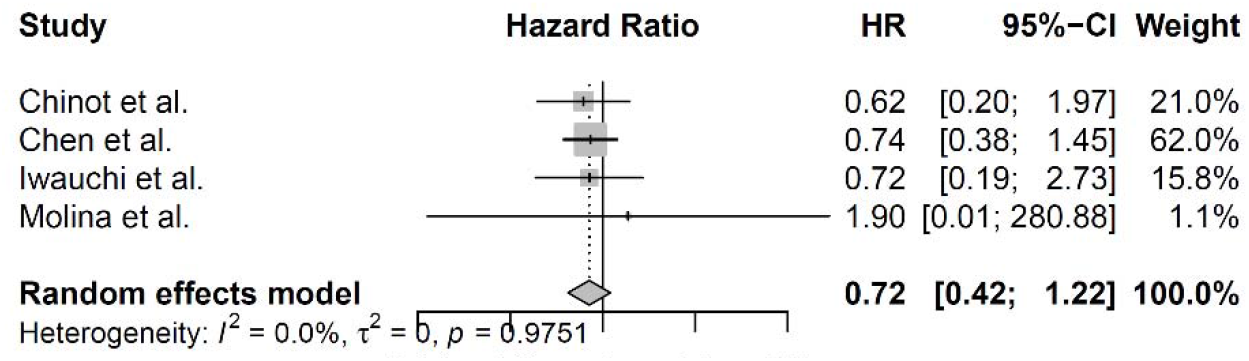
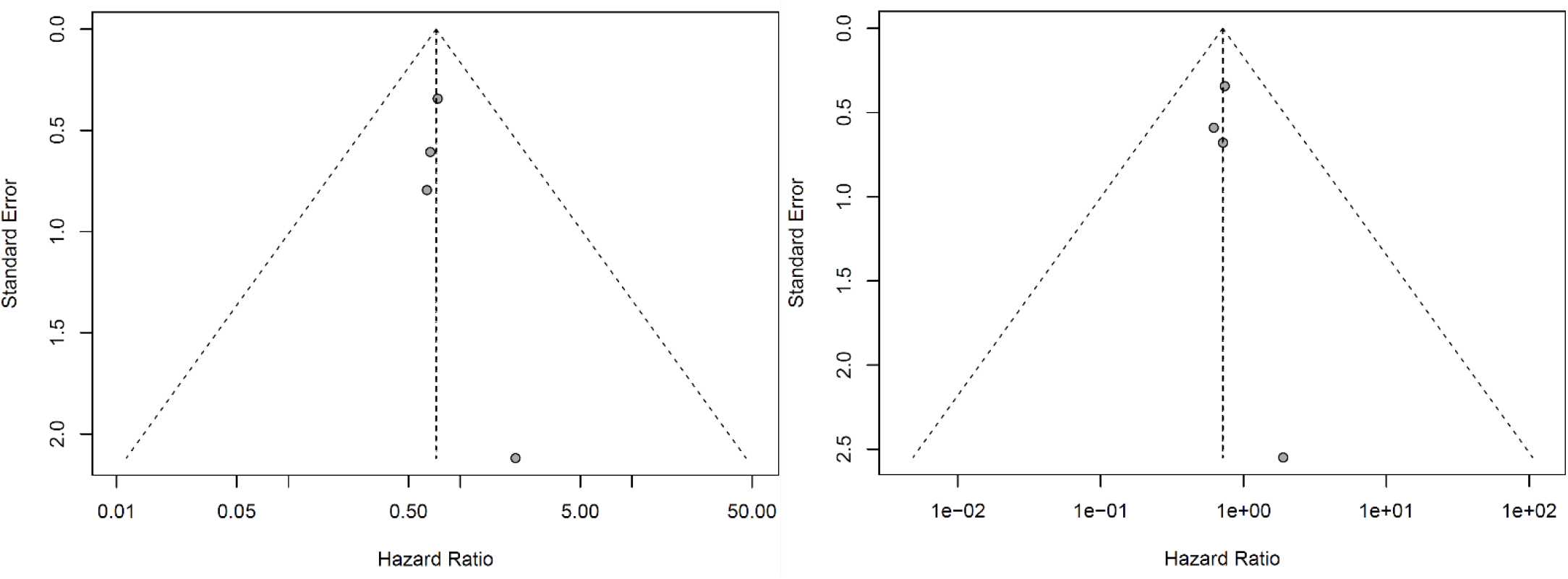
Forest plot of Progression-Free Survival (PFS).

## Discussion

### Summary of Main Findings

In this systematic review and meta-analysis of 10 studies (4 eligible for quantitative survival synthesis; total n = 751), neoadjuvant bevacizumab (BEV, 10 mg/kg per infusion) was evaluated in newly diagnosed, surgically resectable glioblastoma. Across randomized controlled trials (RCTs) and prospective single-arm designs, pooled hazard ratio (HR) estimates showed a numerical, non-significant trend toward benefit in both major endpoints, with minimal statistical heterogeneity. Median OS in smaller non-randomized trials was ≈ 16 months, with PFS ≈ 9 months(13, 27).

### Interpreting PFS–OS Divergence in Bevacizumab-Treated Glioblastoma

Across four studies included in the quantitative synthesis (n = 75 ), neoadjuvant bevacizumab did not demonstrate a statistically significant improvement in either progression-free survival or overall survival in pooled analyses. These findings are consistent with prior trials evaluating bevacizumab in adjuvant and recurrent glioblastoma settings.(12, 13, 17, 23). This PFS–OS divergence mirrors findings from adjuvant and recurrent GBM trials, where anti-VEGF–induced radiographic responses did not consistently translate into prolonged survival(10-12). Biological factors likely contribute, including the highly infiltrative nature of GBM that limits the cytoreductive benefit of vascular normalization, and emergence of adaptive resistance involving hypoxia and alternative angiogenic pathways (20, 21, 27, 29). Interpretation of survival outcomes should also consider the substantial clinical heterogeneity in bevacizumab treatment protocols across studies. Although statistical heterogeneity in the pooled analysis was minimal (I^2^ = 0%), the underlying interventions were not uniform. As shown in Table 3, the number of preoperative BEV cycles ranged from one to three, concomitant therapies varied from BEV monotherapy to BEV combined with temozolomide and radiotherapy, and the interval between the final BEV infusion and surgery ranged from approximately 21 days to more than five weeks. In addition, several studies continued BEV after surgery as maintenance therapy or reintroduced it at recurrence, whereas others used BEV exclusively in the preoperative phase. This variability complicates direct cross-trial comparisons and limits the certainty of pooled outcomes. Consequently, the GRADE certainty of evidence for survival endpoints was downgraded not only for risk of bias and imprecision, but also for indirectness and inconsistency arising from differences in treatment strategies across studies.

### Clinical and Functional Impact

Consistent across RCTs and prospective cohorts, BEV maintained or improved functional scores in the perioperative period(30-32). Several studies reported improvements in functional performance and cognitive measures following neoadjuvant BEV administration, although the magnitude of benefit varied across cohorts and was frequently reported in small or non-randomized datasets.

### Managing Peritumoral Edema: Dexamethasone Versus Bevacizumab

Preoperative BEV induced marked edema reduction by MRI (−54% median/mean in FLAIR volume in Tanaka and Takei studies) (13, 27), which clinically obviated or minimized corticosteroid needs in several trials (12, 13, 20). Compared to dexamethasone, BEV offers a mechanism-driven VEGF blockade reducing vascular permeability without the wide metabolic and endocrine adverse effects of chronic steroids, though cost and access differ substantially(33).

### The Role of Imaging in Evaluating Bevacizumab Efficacy

MRI volumetrics (T1CE, T1Gd, FLAIR) reliably detected tumor and edema shrinkage after BEV.FMISO-PET hypoxia imaging in Suzuki 2023 showed significant hypoxic volume reduction (p = 0.03) but transient effect with recurrence (21). Molina 2016 identified necrotic volume and rim heterogeneity as potential predictive biomarkers for OS benefit, but only in BEV-treated patients (23).Multimodal MRI approaches also supported earlier detection of progression (HERBY pediatric arm) (26).

### Potential implications for radiotherapy planning

Bevacizumab-induced reductions in contrast enhancement and peritumoral edema may also have implications for subsequent radiotherapy planning. By decreasing vascular permeability and reducing FLAIR-defined edema volumes, neoadjuvant BEV could theoretically influence delineation of the gross tumor volume (GTV) and clinical target volume (CTV). However, none of the included studies systematically reported whether these imaging changes resulted in modified radiotherapy target volumes or altered treatment field sizes. Consequently, it remains uncertain whether preoperative BEV exposure leads to smaller radiation fields or changes in radiotherapy planning strategies.From a biological perspective, VEGF inhibition also affects vascular permeability, tumor hypoxia, and microenvironmental dynamics. These mechanisms raise theoretical considerations regarding the risk of late radiation-related toxicities, including radiation necrosis, following subsequent chemoradiotherapy. However, the studies included in this review did not provide sufficient longitudinal toxicity data to determine whether neoadjuvant BEV modifies long-term radiation toxicity profiles.

### Histological Changes and Adaptive Responses

Histopathological analyses indicated transient vascular normalization following bevacizumab exposure, characterized by reduced CD34 expression, decreased ERG signaling, and diminished glomeruloid vascular proliferation. Hypoxia-associated markers such as CA9 and HIF-1α were also reduced in the early post-treatment period. Markers associated with tumor stemness, including nestin and FOXM1, showed decreased expression. In addition, immunological modulation was observed, including reduced PD-L1 expression and regulatory T-cell (Foxp3+) infiltration, together with increased CD8+ cytotoxic T-cell infiltration. However, recurrence specimens demonstrated adaptive resistance patterns, including re-expression of hypoxia markers, up-regulation of VEGF/VEGFR1 signaling, and persistence of perivascular stem-like niches. (17, 20, 21, 27, 28, 34,35 ).

### Potential Biomarkers for Predicting Bevacizumab Benefit

MRI-derived volumetrics (necrotic core size, rim heterogeneity(23)), FMISO-PET hypoxic volume changes (21), and immunohistochemical hypoxia/stemness profiles (21, 23) have emerged as potential predictors. However, none have undergone external validation, and predictive ability remains hypothesis-generating.

### Biological Rationale and Surgical Implications of Neoadjuvant Bevacizumab

The biological rationale for neoadjuvant bevacizumab is based on its anti-angiogenic effects, including vascular normalization, reduction of vascular permeability, and subsequent decrease in peritumoral edema. Across several included studies, radiological analyses demonstrated substantial reductions in edema and contrast-enhancing tumor volume following BEV exposure. For example, Tanaka 2024 reported a mean reduction of 36.2% in contrast-enhancing tumor volume and a 54.0% reduction in FLAIR-defined edema, while Takei 2022 observed comparable median reductions of 38% and 54%, respectively. These radiographic changes may translate into clinically meaningful improvements in neurological function and surgical conditions. Reduction of edema and mass effect may be particularly relevant for tumors located in eloquent brain regions. In such cases, extensive peritumoral edema can obscure tumor–brain interfaces, increase intracranial pressure, and limit the feasibility of maximal safe resection. The functional improvements observed in several included studies, including increases in Karnofsky Performance Status and improved cognitive scores, suggest that preoperative edema control may stabilize neurological status and facilitate safer surgical planning. However, none of the included studies systematically stratified outcomes according to tumor location in eloquent versus non-eloquent regions, and therefore the potential preferential benefit in these subgroups remains speculative. Another important consideration is the interval between diagnosis and surgery required for neoadjuvant therapy. Across included studies, surgery was typically performed between three and five weeks after the final BEV infusion, depending on protocol design. This interval raises theoretical concerns regarding tumor progression in a highly proliferative malignancy such as glioblastoma. However, the available radiological data from Tanaka 2024 and Takei 2022 demonstrated reductions in both enhancing tumor volume and peritumoral edema during the neoadjuvant period, suggesting that tumor burden did not increase during this interval in the reported cohorts. Nevertheless, these findings are derived from aggregate volumetric data, and individual patient response patterns were not consistently reported. Furthermore, it remains unclear whether all studies performed systematic radiological surveillance during the neoadjuvant and washout periods prior to surgery. Interpretation of radiographic responses must also consider the well-recognized phenomenon of bevacizumab-induced “pseudoresponse,” characterized by rapid decreases in contrast enhancement due to reduced vascular permeability rather than true tumor cytoreduction. In several included studies, imaging assessment incorporated both contrast-enhanced T1-weighted sequences and FLAIR imaging to partially address this limitation, as reductions in edema-related signal may reflect vascular effects rather than tumor cell death. However, standardized response assessment frameworks such as RANO criteria were not consistently reported across the included studies, limiting the ability to distinguish true antitumor effects from imaging-based pseudoresponse. Finally, bevacizumab may influence perioperative outcomes through its effects on angiogenesis and wound healing. Quantitative safety data were available from Tanaka 2024 and Chinot 2016. In Tanaka 2024, no intraoperative hemorrhage occurred, although one patient developed grade 3 wound dehiscence with infection. In the randomized trial by Chinot 2016, higher rates of serious adverse events and grade 3–5 toxicities were reported in the BEV arm, including hypertension, thromboembolic events, proteinuria, and delayed wound healing. These findings highlight the potential for perioperative complications and underscore the importance of appropriate washout intervals prior to surgery.

Overall, while the biological rationale for neoadjuvant bevacizumab is supported by radiographic, functional, and molecular observations, the available clinical evidence remains limited. Additional prospective trials incorporating standardized imaging surveillance, RANO-based response assessment, and detailed reporting of surgical outcomes will be necessary to clarify the true surgical value of this strategy.

### Safety of Bevacizumab Administration

Across BEV arms, no intraoperative hemorrhages were reported(36); wound healing issues were infrequent (one grade 3 in Tanaka) (13). In Chinot 2016, grade 3–5 AEs occurred in 72.6% BEV vs 63% placebo, including hypertension, thromboembolic events, and delayed wound healing

### Impact on Surgical Planning

Several surgical series documented improved tumor–brain margin clarity and reduced mass effect after BEV, enabling maximal safe resection and expanding eligibility for awake craniotomy (21, 23). Importantly, 5-ALA fluorescence was not impaired when surgery occurred ≥3 weeks post-BEV(23). These effects, although anecdotal, suggest an underexplored utility in operative strategy.

### Cost-effectiveness Considerations

Chen12020 found BEV + TMZ substantially exceeded willingness-to-pay thresholds due to the absence of durable OS benefit (17). Model sensitivity was driven by drug acquisition cost and PFS–OS disconnect; no neoadjuvant-specific cost-utility study exists.

### Seizure Control and Antiseizure Drug Dependence

Despite the edema-reducing mechanism and evidence from recurrent GBM suggesting reduced seizure burden with BEV (37), none of the neoadjuvant studies systematically reported seizure frequency or ASM dependence. This represents a key evidence gap.

### Additional Observations

Neoadjuvant BEV appeared to enhance surgical planning by improving tumor–brain interface visualization, facilitating safe resection and eligibility for awake craniotomy without impairing 5-ALA fluorescence (13, 38, 39). Economic analysis in Chen 2020 (17)showed incremental cost-effectiveness ratio far above willingness-to-pay thresholds, driven by absent OS gain(37).

### Overall Interpretation

Neoadjuvant BEV in resectable GBM provides consistent radiological and functional gains, transient biological modulation, and possible surgical facilitation, but without demonstrable long-term OS benefit. These findings support its role as a peri-operative adjunct rather than a definitive disease-modifying agent, pending validation in biomarker-enriched, protocol-standardized RCTs with integrated cost-effectiveness and patient-centred endpoints. Evidence from large randomized trials in the adjuvant setting further contextualizes these findings. The AVAGlio and RTOG 0825 trials demonstrated improvements in progression-free survival with bevacizumab when added to standard chemoradiotherapy in newly diagnosed glioblastoma, but effects on overall survival were limited, and impacts on quality of life and functional status were variable. In addition, higher rates of treatment-related toxicity were observed in bevacizumab-treated patients. Because several studies in the present review continued bevacizumab after surgery or at recurrence, these postoperative exposures may represent potential confounders when interpreting survival and functional outcomes attributed to the neoadjuvant phase.

### Integration of evidence certainty

Across outcomes, the certainty of evidence assessed using the GRADE framework was generally low for clinical endpoints such as overall survival, progression-free survival, and functional status, and very low for radiological and biomarker-based outcomes. These ratings were primarily driven by the limited number of randomized trials, small sample sizes, methodological heterogeneity, and reliance on surrogate endpoints in several studies. Accordingly, the findings of this review should be interpreted as hypothesis-generating rather than definitive evidence of clinical benefit. Future trials incorporating standardized protocols, validated biomarkers, and adequately powered randomized designs will be essential to clarify the true therapeutic value of neoadjuvant bevacizumab in glioblastoma.

## Limitation

Several limitations should be considered when interpreting the findings of this review. First, although all included studies administered bevacizumab at a similar dose (10 mg/kg), substantial heterogeneity existed in the number of preoperative cycles, concomitant therapies, and the interval between bevacizumab administration and surgery. In several studies, bevacizumab was also continued postoperatively or at recurrence, making it difficult to isolate the purely neoadjuvant effect of the intervention.

Second, the overall evidence base was limited by small sample sizes and a scarcity of randomized controlled trials. Among the ten included studies, only two were randomized, and the remaining studies were non-randomized trials or observational cohorts. Furthermore, the meta-analysis for overall survival and progression-free survival included only four studies (n = 751), which reduces statistical power and increases susceptibility to type II error.
Third, several outcomes were reported inconsistently across studies. Functional measures such as Karnofsky Performance Status and cognitive outcomes were available only in a subset of studies and were often assessed without blinding or standardized protocols. Radiological and histopathological outcomes were primarily exploratory and relied on surrogate markers with uncertain correlation to long-term survival.

Finally, safety reporting was incomplete. Quantitative adverse-event data were available in only a limited number of studies, and several investigations reported safety outcomes qualitatively or not at all. As a result, the perioperative risk profile of neoadjuvant bevacizumab cannot be fully characterized.

Overall, these limitations contributed to low or very low certainty of evidence for several outcomes according to the GRADE framework and highlight the need for larger, well-designed randomized trials.

## Future Directions

### Given the above limitations and current evidence gaps, several priorities for future research are clear

#### 1. Well-Powered Randomized Controlled Trials

Large, multicenter RCTs with standardized neoadjuvant BEV protocols—fixed timing relative to surgery, defined number of cycles, and predefined criteria for postoperative continuation—are required to robustly evaluate OS, PFS, and patient-centred outcomes.

#### 2. Integration of Translational Endpoints

Prospective correlation of imaging (e.g., volumetric MRI, advanced perfusion, FMISO-PET) with histopathological and molecular markers (hypoxia, stemness, immune modulation) could validate prognostic and predictive biomarkers identified in current studies.

#### 3. Biomarker-Driven Patient Selection

Trials should stratify by baseline MRI necrotic volume, rim heterogeneity, hypoxic burden, and immune profile to identify responder subgroups and avoid unnecessary toxicity in non-responders.

#### 4. Quality-of-Life and Functional Outcomes

Standardized, blinded KPS and neurocognitive assessments, as well as seizure frequency and steroid dependence, should be systematically collected and reported in all neoadjuvant studies.

#### 5. Safety and Perioperative Risk Characterization

Uniform perioperative AE reporting, including wound-healing outcomes, thromboembolic events, and blood pressure control, must be embedded into trial designs to inform surgical safety.

#### 6. Health Economics Analysis

Cost-effectiveness should be evaluated specifically for neoadjuvant BEV, integrating drug costs, hospitalization, functional preservation benefits, and impact on subsequent therapy costs .

#### 7. Combination and Sequencing Strategies

Investigations into neoadjuvant BEV combined with other targeted, immunotherapeutic, or metabolic agents may overcome resistance pathways (e.g., PI3K-mediated hypoxia adaptation) observed in recurrence .

## Conclusion

In patients with newly diagnosed, resectable glioblastoma, neoadjuvant bevacizumab may provide short-term radiographic and functional improvements, particularly through reduction of peritumoral edema and temporary enhancement of surgical conditions. However, pooled analyses did not demonstrate a statistically significant improvement in overall survival, and the certainty of the available evidence remains low to very low according to the GRADE framework. Current evidence is therefore insufficient to support routine clinical adoption of neoadjuvant bevacizumab outside research settings. Its potential perioperative utility, especially for edema control, functional stabilization, and surgical facilitation, should be further investigated in well-designed randomized trials with standardized treatment protocols and biomarker-guided patient selection.

## Data Availability

All data produced in the present study are available upon reasonable request to the authors

## Declaration of AI Use

Artificial intelligence (AI) tools (ChatGPT) were used to improve the grammar, clarity, and language of this manuscript. The authors reviewed and approved all content, and take full responsibility for the integrity and accuracy of the work.

## Acknowledgments

The authors have no acknowledgments to declare.

## Appendix and Supplementary Material

- Appendix 1. Systematic Review Protocol
- Appendix 2. Search Strategy
- Appendix 3.Certainty of evidence (GRADE approach)
- Appendix 4. Data extraction Sheets
- Appendix 5. Risk of bias Tables
- Appendix 6. Excluded studies

